# Evaluation of Clinical Job Demands, Job Resources, and a Novel Intervention on Measures of Health Care Worker Stress at a Community Hospital Pre and Post COVID-19

**DOI:** 10.1101/2021.07.06.21260093

**Authors:** Jessica Sels, Dylan Carroll, Doug Duffee

## Abstract

**Objective:** To explore the relationship and identity of Health Care Worker stressors to a measure of perceived burnout and to a novel intervention tool.

**Participants and Methods:** From July 2019 to June 2020, we surveyed Health Care Workers (HCW) pre and post COVID19 in an independent local community hospital for burnout with the Health Care Provider Wellness Assessment tool. Linear regression and means comparison were used to identify overall job demand and resource perception with burnout, unique stressor portraits by provider subtype and mean survey scores between those who did or did not voluntarily complete at least 14 days of a 28 day novel self-help intervention tool.

**Results:** Regarding the pre COVID-19 data, of 73 respondents, there was statistically significant (p<.01) correlation between overall job demands (directly) and resources (inversely) with burnout intensity. With respect to the HCW stressor characteristic analyses there was statistical significance (p<.05) between the mean frequency occurrence of the top 5 stressors identified by respondent subtype when compared to the mean occurrence of overall individual responses within the corresponding subtype. Finally, although limited by a low number of respondents, the intervention tool analysis suggested a therapeutic trend toward disruption of the stress-burnout relationship. Regarding the post COVID-19 data, 18 respondents did not show statistically significant characterizable stressor portraits (ie stressors were present but not patternable).

**Conclusion:** Unique stressor portraits were identified by HCW subtype which correlated with more intense burnout self-perception. Additionally, there was a trend toward self-help tool efficacy in mitigating burnout.

## Introduction

The problem of health care worker burnout has been well documented (1-4). As health care workers encounter the demands and resources of a rapidly changing health care system and navigate their place and performance within it, deal with the demands of an internet informed patient populace and balance workload with family life, stressors arise. These stressors can contribute to burnout and this burnout has physiologic effects (5,6,7,10,22) and predicts objective associations with health care acquired infection rates, medical errors, medical litigation, patient satisfaction, job satisfaction, health care system costs, alcohol abuse and suicidal ideation, among others(4). As our population ages and its medical co-morbidities and system demands increase (8), the premature curtailing, cessation or turnover of an HCW’s clinical practice due to emotional exhaustion is a concerning trend (9). While the data is mixed as to whether or not HCW’s are more or less prone to burnout than other professionals(4), a growing body of literature has shown that an intentional focus on institutional processes that nurture clinician well-being through multiple modalities is both important(2) and effective(11,19). Thus identifying both the stressed and the stressors becomes paramount. Additionally, insights into the role that self-help tools incorporating forgiveness play in personal well-being (12-17) prompt our presentation as well of a novel mindfulness activity focusing on HCW wellbeing by incorporating personal forgiveness exercises. Also in light of recent studies that have cautioned against the tendency to dichotomize and/or pathologize (3) peoples’ responses to their work environment, we used the JD-R (Job demands-resources) model and the CBI (Copenhagen Burnout Inventory) presented as the components of our HCPWA (Health Care provider Wellness Assessment) survey to capture workplace dynamics and individual coping responses. These factors have prompted us to present a study designed to evaluate wellness affecters inside the culture of an independent community teaching hospital and to evaluate the effectiveness of an intervention which aims at voluntary facilitated self-help with an eye on both for wider application.

### Participants and Methods

From July 2019 to February 2020 and again from April 2020-June 2020 healthcare workers at Parkview Medical Center (an independent medical hospital and medical group in Pueblo, Colorado) and associated clinics were asked to complete the HCPWA, a compilation of the JD-R(22) and CBI(20). The survey was accessed through an independent website and aggregated by Survey Monkey. Respondents identifying as Medical Assistants, RN’s and Providers (Nurse Practitioners, Physician Assistants, Physicians or Pharmacists) voluntarily participated in response to a general solicitation for participation. A total of 91 healthcare providers responded (73 pre-Covid19 and 18 post-Covid19). Linear regression analysis was performed to evaluate for correlation between job demand-resource stress categories and overall burnout score intensity. Additionally means testing was performed to identify unique stressor characteristics and to assess disruption to a stress-burnout relationship by an intervention tool.

### Survey

The survey, called the Health Care Provider Wellness Assessment (HCPWA), is a 71 question wellness assessment tool derived from our combination of the modified Job Demands-Resource (JD-R) work stress model and the Copenhagen Burnout Inventory (CBI). The combination of these 2 assessment tools into one survey was unique to this study and sought to leverage the strength of the JD-R in quantifying environmental stressors coupled with the strength of the CBI in quantifying individual subject burnout response to said stressors inside the context of professional activity. Both tools are available for use in the public domain.

The JD-R, designed to quantify workplace stressors is built as an “overarching model that may be applied to various occupational settings irrespective of particular demands and resources…based on underlying psychological processes” (22). The modification of the JD-R in 2004 sought also to capture work engagement characteristics (as mitigating criteria to burnout)(23). We phrased the survey questions to capture each of the demands and resources stressors as listed as part of the model (23).

In light of recent recommendations on the utility of the CBI as well as the near universal use of the MBI being called into question(3), we elected to use the CBI as the quantitative measure of burnout in this study. The CBI is a 19 question survey covering 3 components of burnout defined as personal, work and client related. These domains are designed to assess burnout universally, inside professional activity and inside the professional therapeutic context. For purposes of the CBI, the term client is understood in the appropriate context as “patient” (as opposed to customer or colleague). There is high internal reliability of the CBI, the questions are easy to understand and answer and it has been shown to correlate well with SF-36(24). In this study we used the CBI in 2 ways, initially as described under statistical analysis, we broke down the linear regression by the average score of individual questions. We used the total burnout score (>= 75 to a max of 95) to identify a subtype of respondents that were “burnt out”.

Except for 2 initial demographic questions, the 69 environment stressor and burnout characteristic questions (50 stressor and 19 burnout questions) were answered by respondents on an intensity scale from 1-5.

### Intervention Tool

The intervention tool was designed as a self help tool consisting of a daily 5 minute introspective reflection on an ancient semitic poem called Psalm 19. The facilitated study guide called “Psalm 19 Insights and Exercises in Personal Forgiveness” explored topics such as perfectionism, loneliness, prayer, fear and guilt and others designed to be completed over 28 days. Respondents were offered the tool to be voluntarily completed prior to taking the survey.

### Statistical Analysis

The full survey comprised 69 items capturing 24 job demand stressors, 26 job resource stressors and 19 items exploring an individual’s self-perception of burnout. Each is measured on a five-point attribution scale (1=never/almost never, 2=seldom, 3=sometimes, 4= often, 5=always), so the theoretical range is 1-5.

Initial data analysis involved looking for overall correlation between job demands and resources with burnout. In order to refine the data for analysis, composite measures of each variable are created by summing and averaging the constituent elements. The overall results are shown in Table 1. Burnout scores range from 1.26 to 4.74, with a mean score of 3.1, indicating an average burnout score. Mean scores for job resources (3.0) and job demands (3.2) also fall in the middle of the range indicating that most participants selected ‘sometimes’ for most of the items comprising the variables.

**Table 1:**
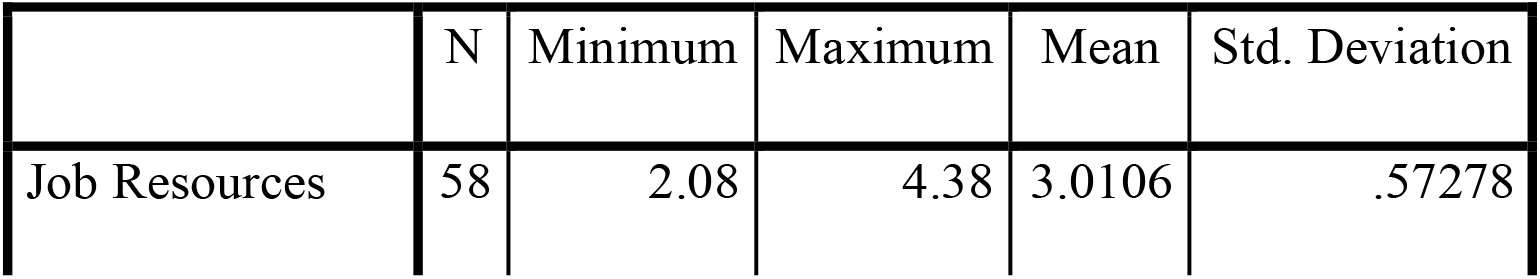
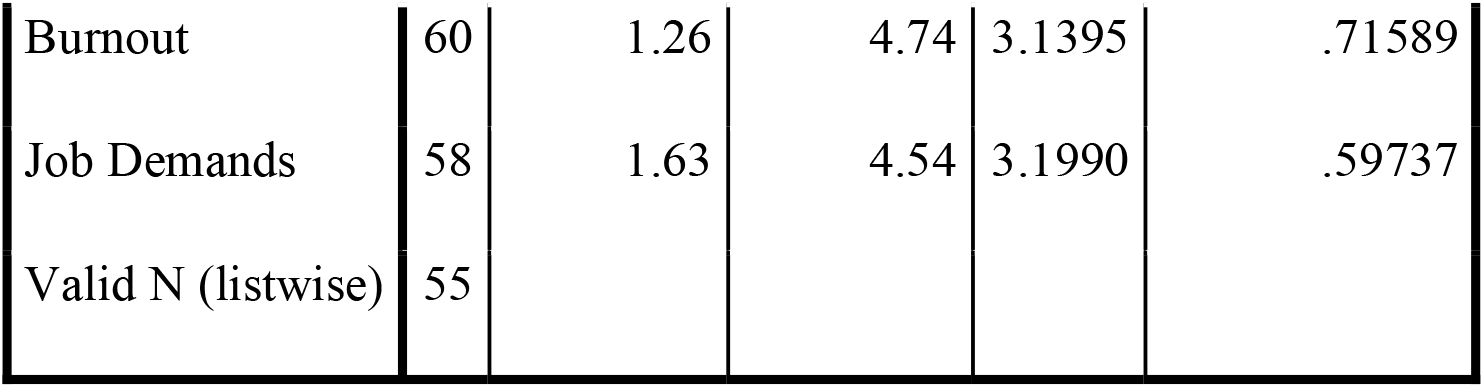
Summary Results, JDR Model.

Table 2 demonstrates that all three pairs of variables are correlated with one another in the intended direction. Specifically, there is a strong, positive relationship between job demands and burnout such that as job demands increase, so do perceptions of burnout and strain. This relationship is depicted visually in Figure 2. In addition, there is a strong, negative relationship between job resources and burnout such that as job resources increase, perceptions of burnout and strain decrease (Figure 3).

**Table 2:**
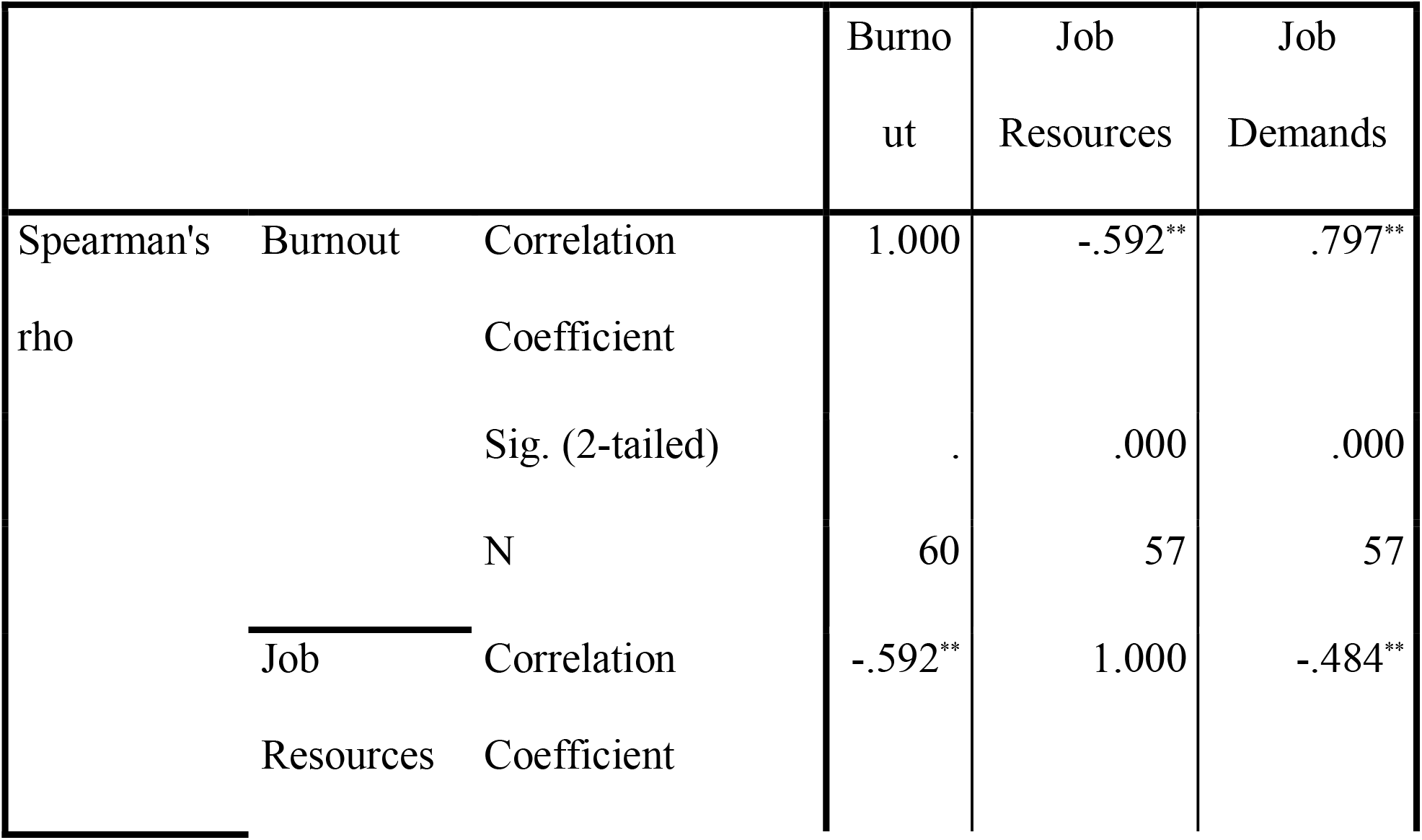
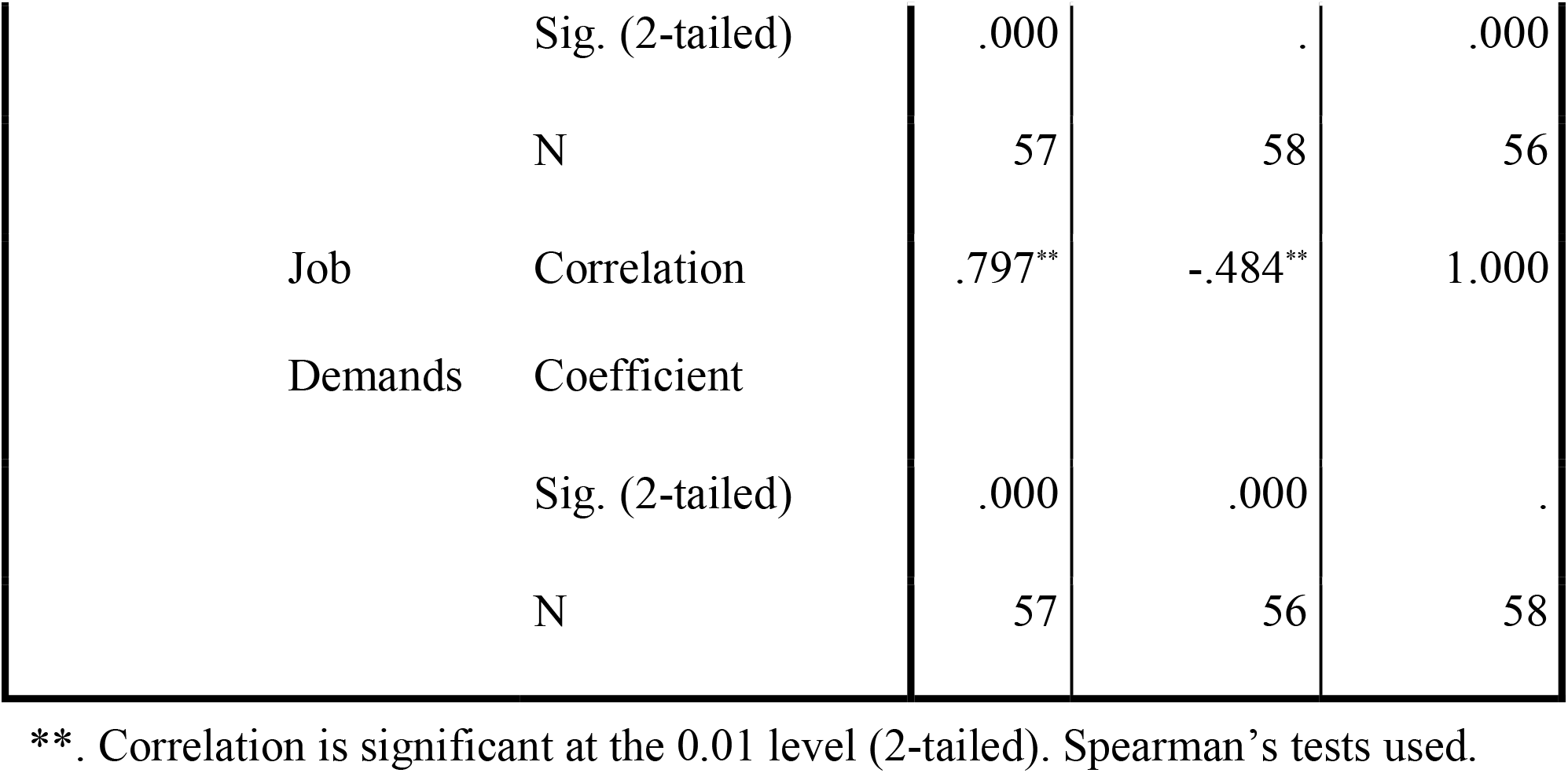
Intercorrelations, JDR Model.

**Figure 1:**
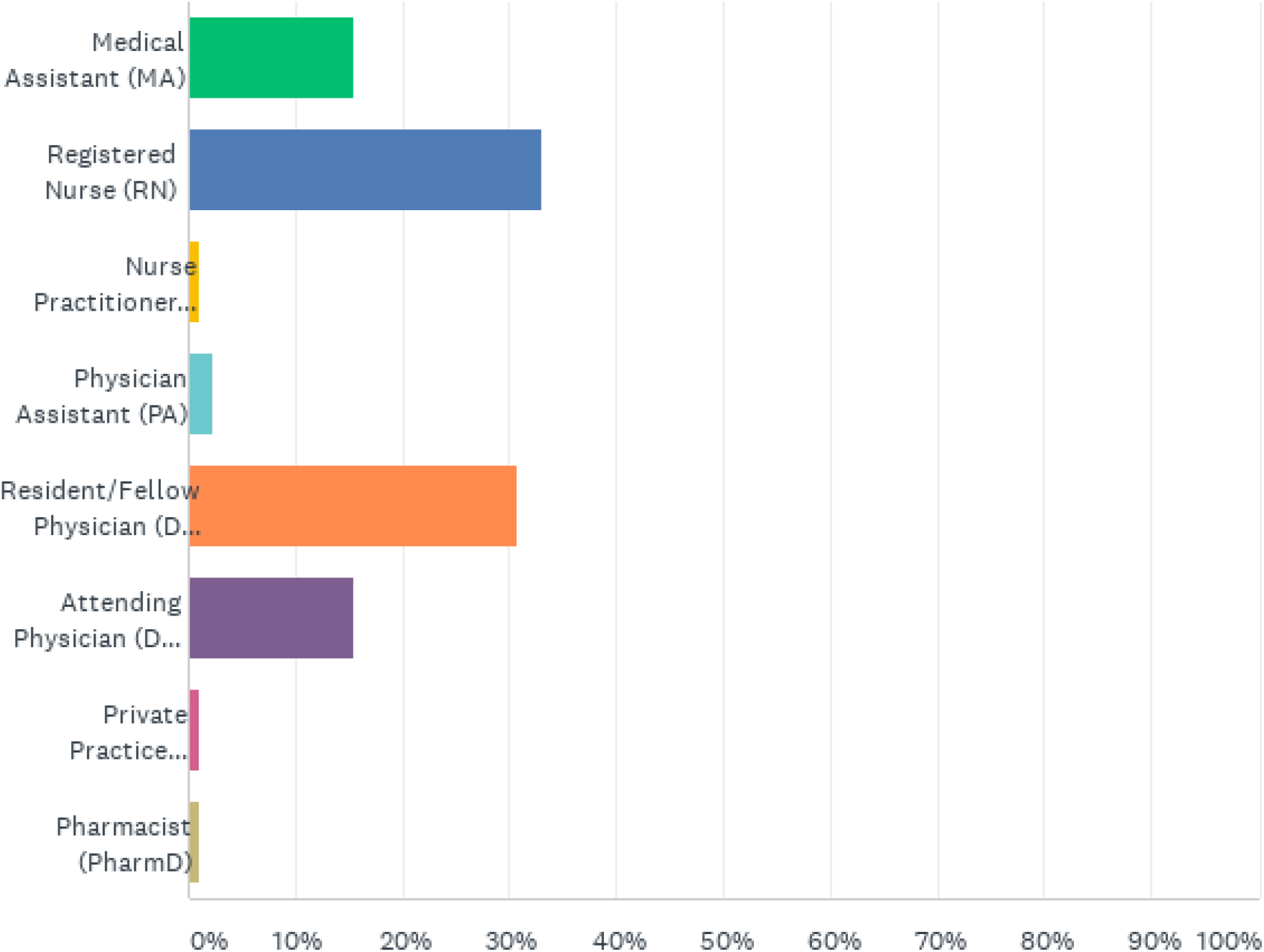
Role of the Respondents.

**Figure 2:**
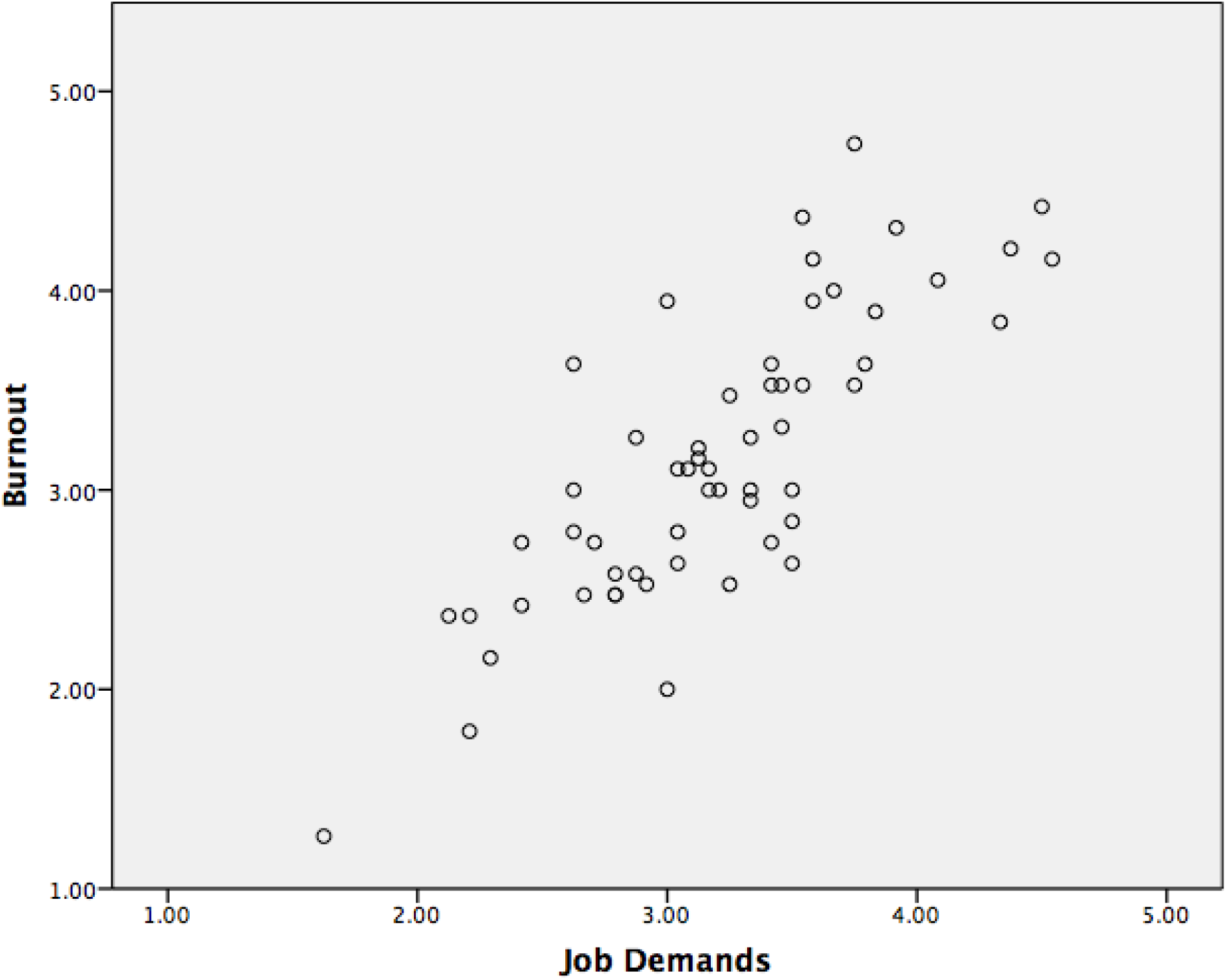
Relationship between job demands and burnout.

**Figure 3:**
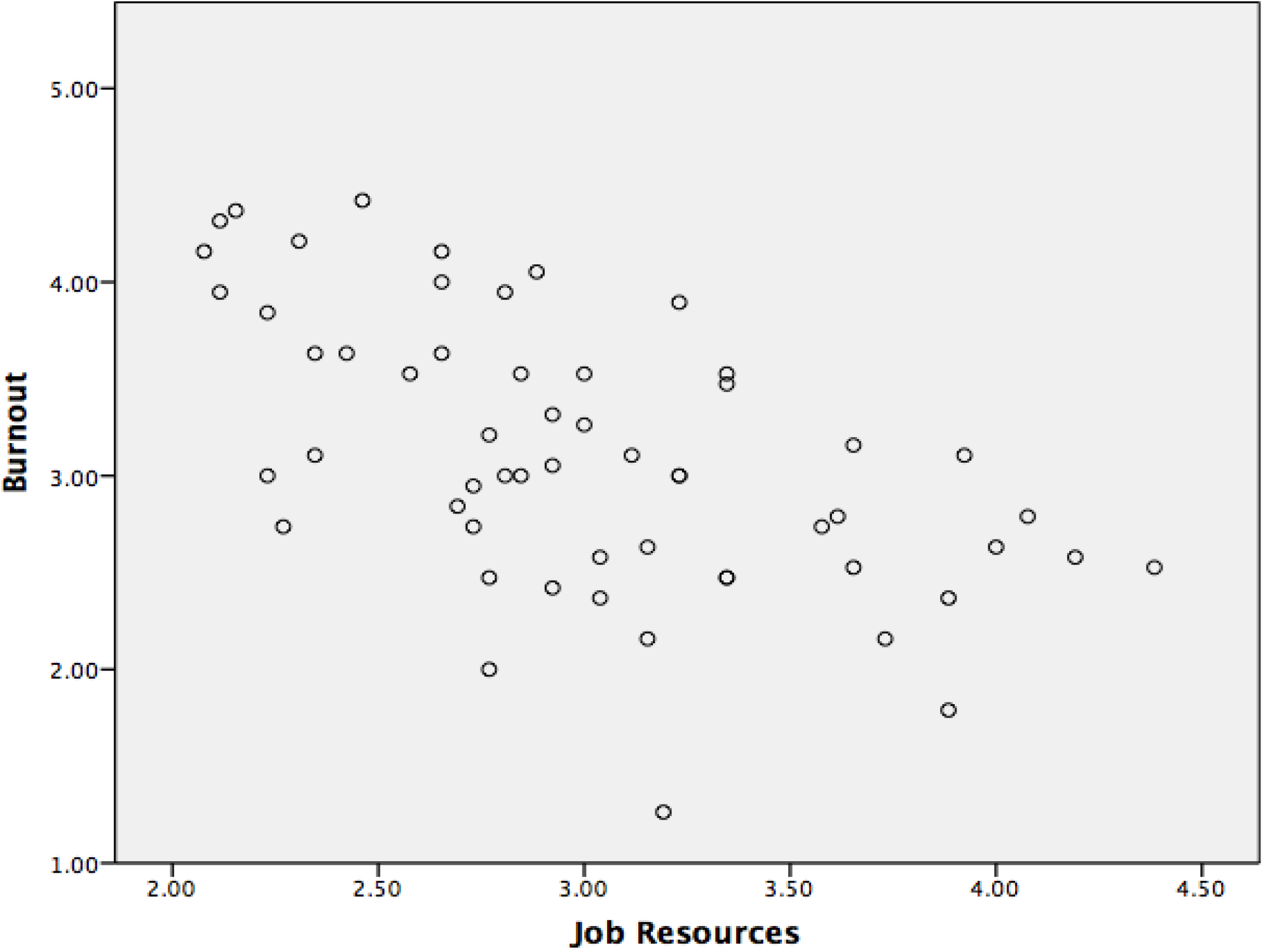
Relationship between job resources and burnout.

The above results are confirmed in a linear regression analysis (Table 3), which demonstrates that job demands significantly increase burnout, and job resources significantly reduce perceptions of burnout.

**Table 3:**
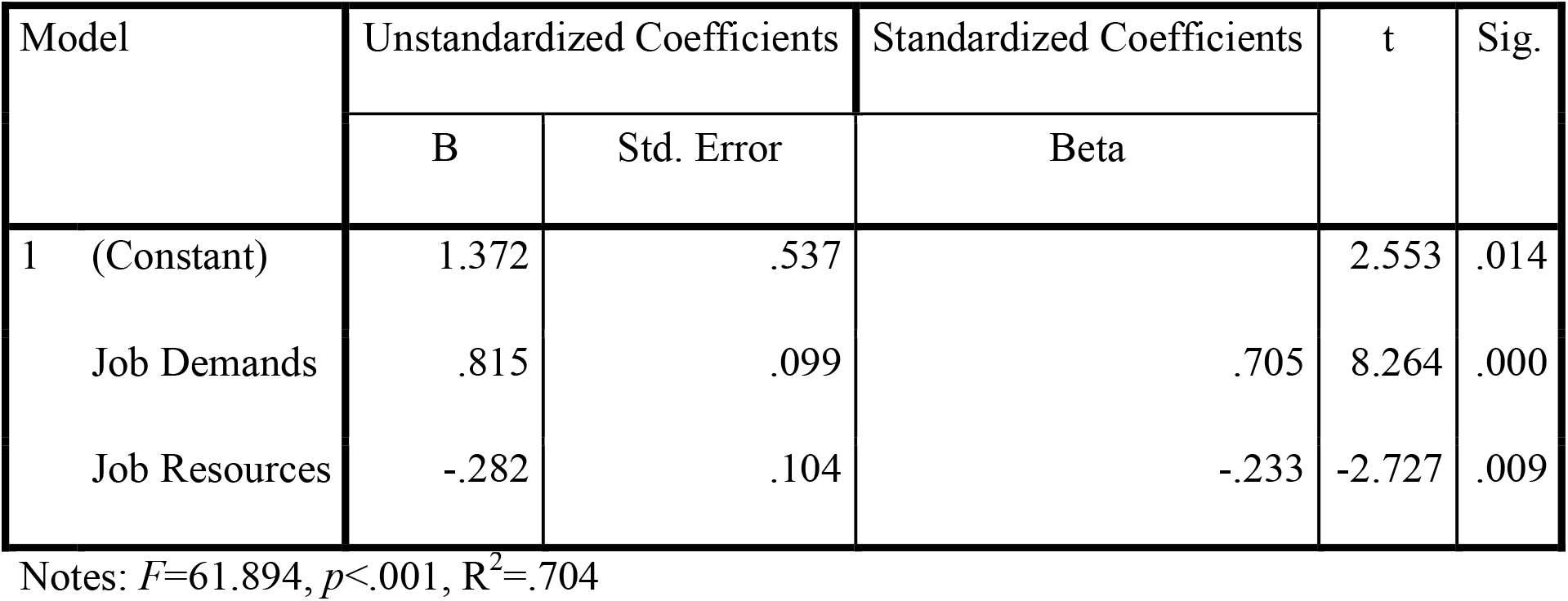
Linear Regression Analysis, JDR Model.

| Model | Unstandardized Coefficients |  | Standardized Coefficients | t | Sig. |
| --- | --- | --- | --- | --- | --- |
|  | B | Std. Error | Beta |  |  |
| 1 (Constant) | 1.372 | .537 |  | 2.553 | .014 |
| Job Demands | .815 | .099 | .705 | 8.264 | .000 |
| Job Resources | -.282 | .104 | -.233 | -2.727 | .009 |

Notes: $F=61.894$ , $p<.001$ , $R^2=.704$

Subsequent analysis evaluated the impact of the wellness intervention on burnout. It is theorized that the completion of the wellness intervention will disrupt the observed relationship between job demands and burnout, enabling participants to make better use of their existing job resources. In the first instance, the dataset is disaggregated by those who completed less than 14 days of the intervention, and those who have completed more of the intervention. Mean burnout levels are expected to differ significantly between the two groups. Indeed, the results show that individuals who have completed fewer than 14 days of the intervention perceive significantly higher burnout and higher environmental stress (higher job demands, less resources) than individuals who have completed more than 14 days of the intervention.

The small sample sizes for those that have completed 14 days or more of the intervention do make performing correlation analysis statistically challenging. A regression analysis is performed that disaggregates the dataset according to completion of the tool, and while the F value for those that had completed 14 days or more (*n*=5) indicates poor model fit, it can be seen that the statistically significant relationship between job demands, job resources and burnout observed earlier disappears once the tool has been completed. In other words, there is preliminary evidence that completing at least 14 days of the intervention tool prevents high job demands and low job resources being translated into burnout.

Final analysis involved a means comparison between the top 5 demand and resource items which garnered the most high intensity (level 5) responses. The mean of these top 5 grouped responses were compared to the mean number of overall high intensity responses per survey item inside each subtype to assess whether these stressors were more commonly perceived compared to the others. The “top 5” were chosen (as opposed to more or less) in order to provide a reasonable amount of actionable items. This was accomplished through the use of a Wilcoxon Signed-Rank Test Calculator pre and post Covid analysis.

Results are summarized in the table 6 as stressor portraits by provider subtype:

**Table 4:** Comparison of Mean JDR Model.

|  |  | N | Minimum | Maximum | Mean | Std.<br>Deviation |
| --- | --- | --- | --- | --- | --- | --- |
| Completed less than 14 days | Job Resources* | 51 | 2.08 | 4.38 | 2.9532 | .56925 |
|  | Burnout** | 53 | 1.26 | 4.74 | 3.2592 | .66964 |
|  | Job Demands** | 52 | 1.63 | 4.54 | 3.2796 | .56702 |
|  | Valid N (listwise) | 49 |  |  |  |  |
| Completed 14 days or more | Job Resources | 5 | 3.04 | 3.88 | 3.4692 | .33794 |
|  | Burnout | 5 | 2.16 | 2.47 | 2.3684 | .12892 |
|  | Job Demands | 4 | 2.13 | 2.79 | 2.4479 | .33050 |
|  | Valid N (listwise) | 4 |  |  |  |  |

Comparison undertaken using Mann Whitney U tests. \* $p < .001$ , \*\* $p < .005$ ,

**Table 5:** Linear Regression Analysis, JDR Model, by Completion of the Wellness Intervention Tool.

| Model |  | Unstandardized Coefficients |  | Standardized Coefficients | t | Sig. |
| --- | --- | --- | --- | --- | --- | --- |
|  |  | B | Std. Error | Beta |  |  |
| Completed less than 14 days | 1 (Constant) | 1.525 | .541 |  | 2.819 | .007 |
|  | Job Resources | -.297 | .103 | -.260 | -2.885 | .006 |
|  | Job Demands | .789 | .103 | .690 | 7.658 | .000 |
| Completed 14 days or more | 1 (Constant) | 2.046 | .118 |  | 17.348 | .037 |
|  | Job Resources | -.018 | .026 | -.107 | -.698 | .612 |
|  | Job Demands | .179 | .028 | .974 | 6.375 | .099 |

Notes: Model 1 $F=51.188$ $p<.001$ , $R^2=.690$ ; Model 2 $F=21/095$ $p=.152$ , $R^2=.977$

**Table 6:**
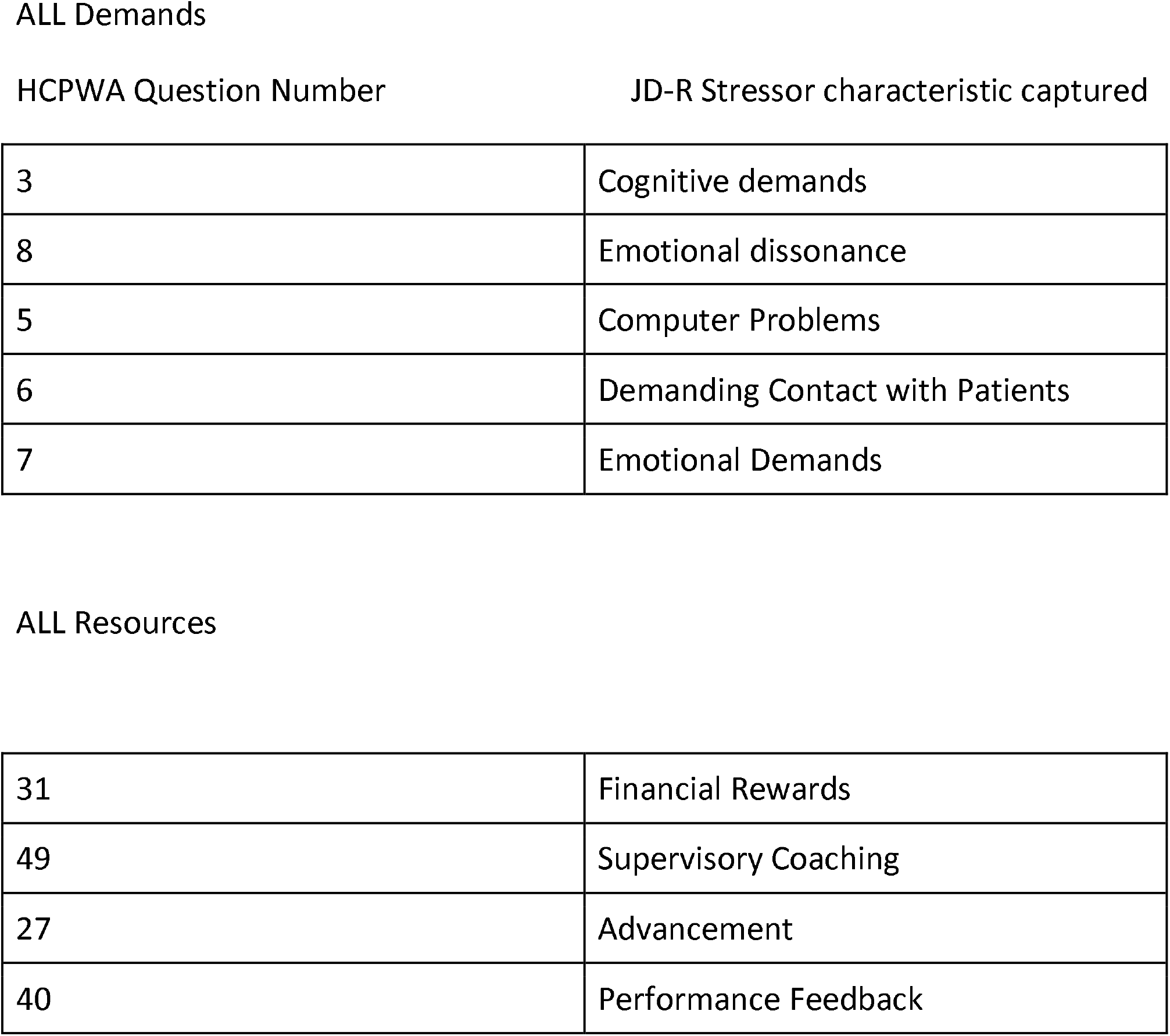
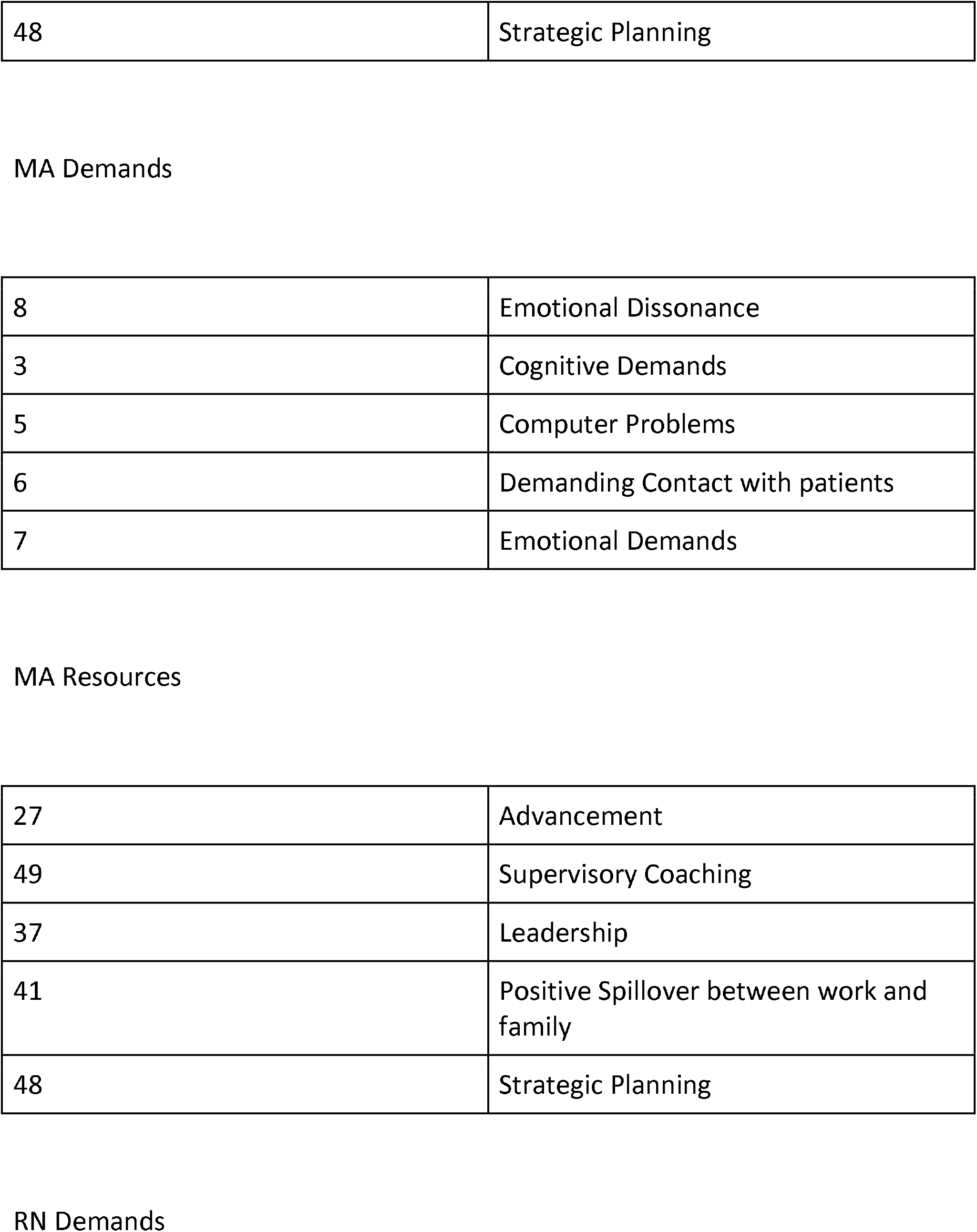
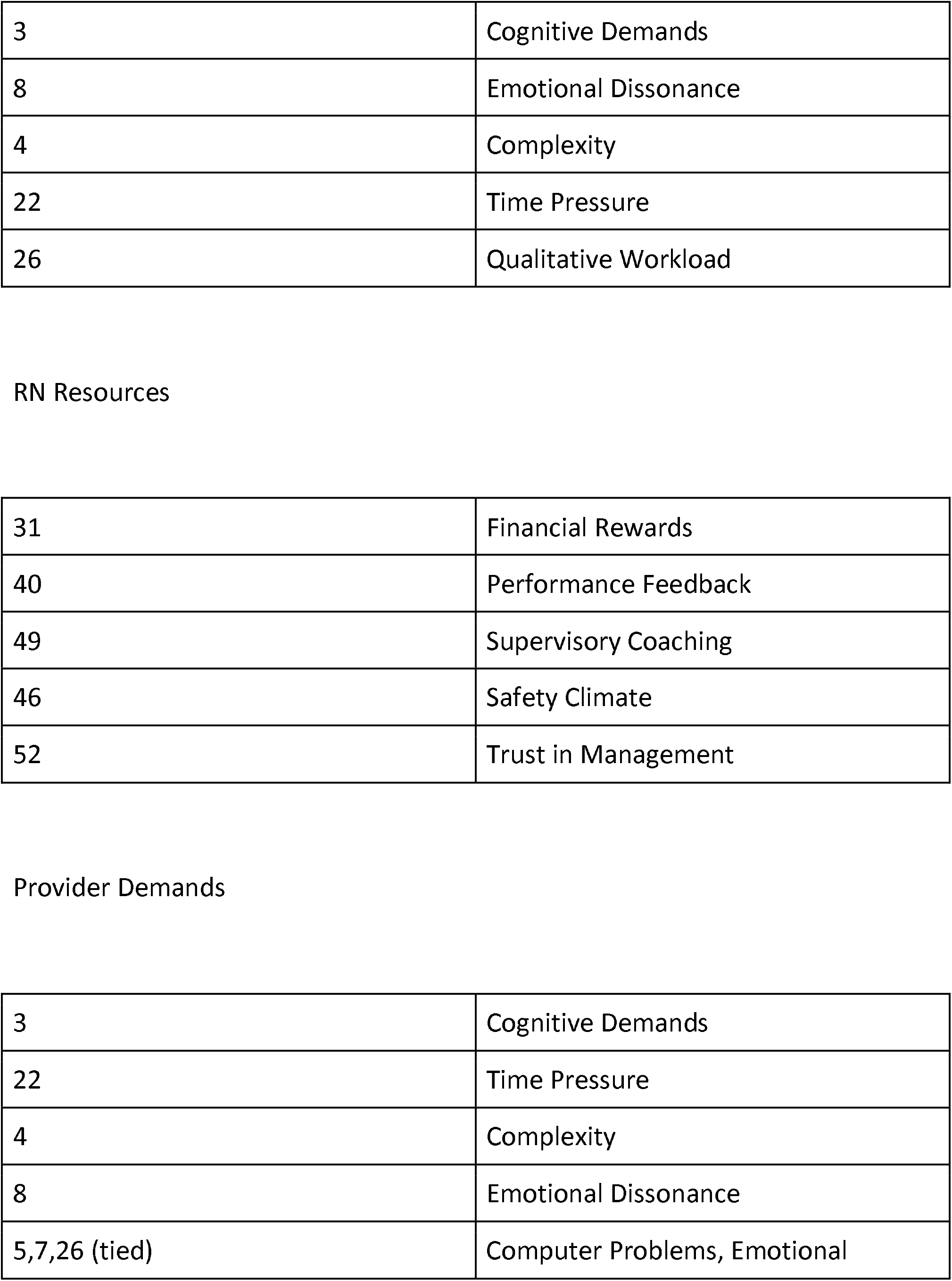
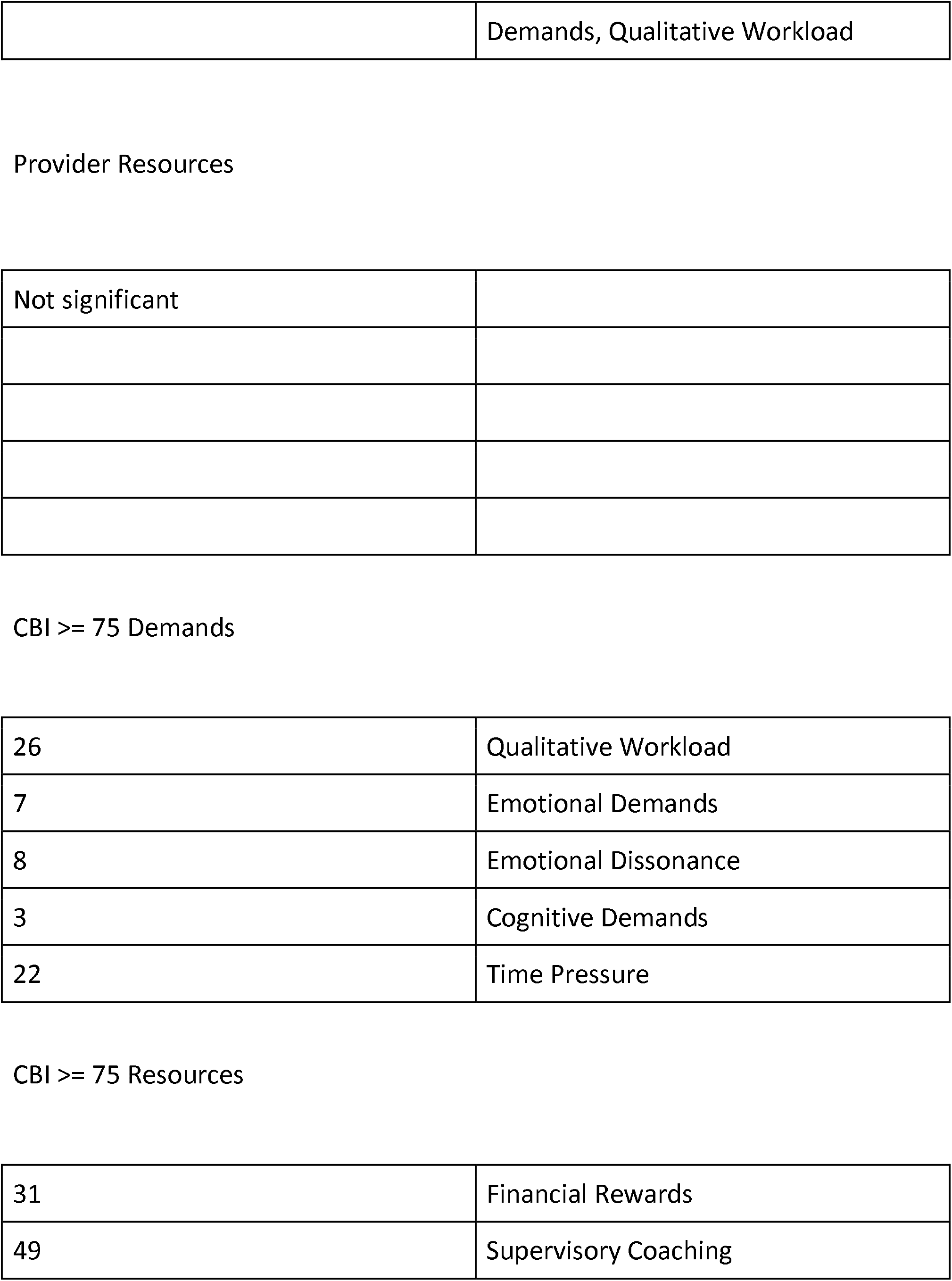
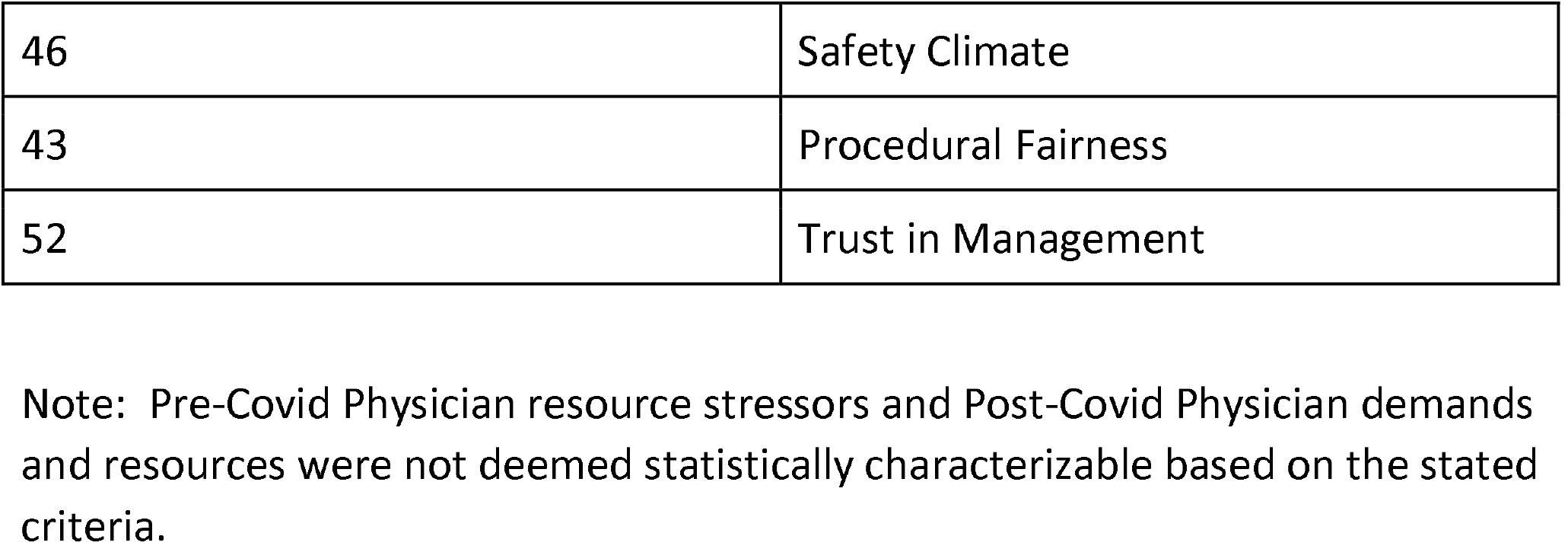
Stressor Portraits by Provider Subtype.

|  |  |
| --- | --- |
| 3 | Cognitive demands |
| 8 | Emotional dissonance |
| 5 | Computer Problems |
| 6 | Demanding Contact with Patients |
| 7 | Emotional Demands |
| ALL Resources |  |
| 31 | Financial Rewards |
| 49 | Supervisory Coaching |
| 27 | Advancement |
| 40 | Performance Feedback |
| 48 | Strategic Planning |
| MA Demands |  |
| 8 | Emotional Dissonance |
| 3 | Cognitive Demands |
| 5 | Computer Problems |
| 6 | Demanding Contact with patients |
| 7 | Emotional Demands |
| MA Resources |  |
| 27 | Advancement |
| 49 | Supervisory Coaching |
| 37 | Leadership |
| 41 | Positive Spillover between work and family |
| 48 | Strategic Planning |
| RN Demands |  |
| 3 | Cognitive Demands |
| 8 | Emotional Dissonance |
| 4 | Complexity |
| 22 | Time Pressure |
| 26 | Qualitative Workload |
| RN Resources |  |
| 31 | Financial Rewards |
| 40 | Performance Feedback |
| 49 | Supervisory Coaching |
| 46 | Safety Climate |
| 52 | Trust in Management |
| Provider Demands |  |
| 3 | Cognitive Demands |
| 22 | Time Pressure |
| 4 | Complexity |
| 8 | Emotional Dissonance |
| 5,7,26 (tied) | Computer Problems, Emotional |
|  | Demands, Qualitative Workload |

Provider Resources
|  |
| --- |
| Not significant |
| CBI >= 75 Demands |  |
| 26 | Qualitative Workload |
| 7 | Emotional Demands |
| 8 | Emotional Dissonance |
| 3 | Cognitive Demands |
| 22 | Time Pressure |
| CBI >= 75 Resources |  |
| 31 | Financial Rewards |
| 49 | Supervisory Coaching |
| 46 | Safety Climate |
| 43 | Procedural Fairness |
| 52 | Trust in Management |

Note: Pre-Covid Physician resource stressors and Post-Covid Physician demands and resources were not deemed statistically characterizable based on the stated criteria.

Additionally, ANOVA calculations showed no statistically significant difference in CBI scores between provider subtype.

## Results

Of the 91 health care providers surveyed pre and post covid, 87% completed the survey and were included in the analysis. The demographic makeup of the study group is illustrated in Table 1. 15.38% were MA (14), 32.97% RN (30), 1.10% NP (1), 2.20% PA (2), 30.77% resident/fellow (28), 15.38% attending physician (14), 1.10% private practice physician (1), 1.10% pharmacist (1). Initially, HCPWA response scores were averaged in demand, resource and burnout categories and compared for correlation as outlined in Tables 1 and 2 and Figures 1 and 2. There was strong correlation between the demand and resource stressor categories and a participant’s self-perception of burnout. While this was expected, the strong numerical significance provided support for the means approach to analyzing stress and burnout as captured by the HCPWA. Thus the 5 component stressor portrait was obtained by HCW subtype via Wilcoxen means comparison and except as noted in the statistical analysis discussion, was able to provide the “group of 5” stressors that were significantly more stressful to participants than the average of intensity responses for the other questions. We also used the overall score on the CBI to identify a separate subgroup of health care providers that meet the intensity criteria for burnout (>=75-95 on the 19 question CBI).

While the intervention tool seemed to disrupt the stressor burnout relationship, the statistical analysis was complicated by low numbers of participants who completed >= 14 days of it.

## Discussion

In this cross sectional survey of Health Care workers in a community hospital pre and post-Covid19 presentation, utilization of the HCPWA was able to confirm associations between increased job demands, decreased job resources and burnout intensity. This association was quantified in a way that used the juxtaposition and combination of 2 well validated wellness tools in order to more specifically and standardly clarify what particular components of stress were being experienced by subtypes of HCW’s as well as those identifying as “burnt out”. Additionally the illustration of stressor portrait subtypes allows a clearer understanding of the burnout experience by department. Using means comparison of a group of 5 stressors to overall stressors allows for more specific identification of and intervention inside the burnout experience. In that the wellness intervention tool suggested an ability to disrupt the stressor-burnout relationship, utilization of a self-help wellness exercise inside burnout intervention may be a reasonable tool to study further for use in the wellness tool belt. As a model for work environment stress and burnout assessment, the use of the HCPWA may be generalizable to other health care institutions and contexts.

This study had several limitations. First, it was conducted at a single organization on a voluntary basis that may have contributed to selection bias both inside the survey and the intervention tool. Secondly, as noted in the statistical analysis, the number of study participants completing at least 14 days of the intervention tool was small. Also, while the HCPWA survey was written in simple language, perception of the issue of concern behind the question may have been understood differently between participants for some questions.

## Conclusion

In this cross sectional survey of health care workers in the community medical practice setting, the Health Care Provider Wellness Assessment and a self-help wellness intervention tool were able to confirm and more specifically identify a strong correlation between professional job demands, resources and burnout intensity and to disrupt the relationship between them.

## Data Availability

All raw/original data XLS is stored on secure institutional drive

https://www.surveymonkey.com/r/SLD5NTT

## Abbreviations and Acronyms

HCW: Health Care Worker
HCPWA: Health Care Provider Wellness Assessment
JD-R: Job Demands and Resources
CBI: Copenhagen Burnout Inventory
MBI: Maslach Burnout Inventory

## Grant Support

The work was funded by the Department of Internal Medicine and Graduate Medical Education, Parkview Medical Center. Funding sources had no role in study design; in the collection, analysis, and interpretation of data; in writing of the report; and in the decision to submit the article for publication.

## Potential Competing Interests

The authors report no competing interests

## Notes

### Competing Interest Statement

The authors have declared no competing interest.

### Clinical Trial

NCT04129632

### Funding Statement

None

### Author Declarations

Protocol was approved by Parkview Medical Center IRB

## Bibliography

1. Yaribeygi H, et al., The impact of stress on body function: A review. EXCLI Journal. 2017 Jul 21; 16: 1057-1072

2. Shanafelt, Tait D. et al., Executive Leadership and Physician Well-being, Mayo Clinic Proceedings January 2017, 92, 1, 129-46

3. Rotenstein, Lisa S., Prevalence of Burnout Among Physicians, JAMA 2018; 320(11), 1131–1150.

4. Dyrbye et al., Burnout Among Health Care Professionals, A Call to Explore and Address This Under recognized Threat to Safe, High-Quality Care. NAM Perspectives. Discussion Paper, National Academy of Medicine.

5. Henry, James P., Biological Basis of the Stress Response, Integrative Physical and Behavioral Sciences, January 1992, Vol. 27, 1, 66–83.

6. McEwen, Bruce S., Neurobiologic and Systemic Effects of Chronic Stress, 2017, Chronic Stress.

7. McEwen Bruce S., Protective and Damaging Effects of Stress Mediators, NEJM 338; 171–179.

8. Health, US report, 2016, CDC.

9. Silver MP, et al., A systematic review of physician retirement planning. Human Resources for Health. 2016 Nov 15; 14: 67

10. Julkanen, J. et al., Hostility and the progression of Carotid Atherosclerosis, Psychosomatic Medicine, 56, 519–525.

11. Cavanagh K, Strauss C, Forder L, Jones F. Can mindfulness and acceptance be learnt by self-helpã: a systematic review and meta-analysis of mindfulness and acceptance-based self-help interventions. Clin Psychol Rev. 2014 Mar;34(2):118–29. doi: 10.1016/j.cpr.2014.01.001. Epub 2014 Jan 10. Review.

12. Harris, Alex HS., et al, Forgiveness, Unforgiveness, Health and Disease from Handbook of Forgiveness 2005, Routledge, Everett L. Worthington, ed.

13. Thoresen, CE., et al, Effects of Forgiveness Intervention on perceived stress, state and trait anger and self-reported health. Paper presented at annual meeting for society of behavioral health, Seattle WA, 2001.

14. Waltman, MA., The psychological and physiological effects of forgiveness education in male patients with coronary artery disease. Dissertation Abstracts International: Section B: The Sciences and Engineering; 63(8-B), 3971.

15. Witvleit, C., forgiveness and Health: Review and reflections on a matter of faith, feelings and physiology, Journal of Psychology and Theology, 29, 212–224.

16. Witvleit, C., et al., Please forgive me: Transgressors’ emotions and physiology during imagery of seeking forgiveness and victim responses. Journal of Psychology and Christianity, 21, 219–233.

17. Witvleit, C., et al, Granting forgiveness or harboring grudges: Implications for emotion, physiology and health, Psychological Science, 12, 117–123.

18. Worthington EL Jr, Witvliet CV, Pietrini P, Miller AJ. Forgiveness, health, and well-being: a review of evidence for emotional versus decisional forgiveness, dispositional forgivingness, and reduced unforgiveness. J Behav Med. 2007 Aug;30(4):291-302. Epub 2007 Apr 24. Review.

19. Lupano Perugini ML, de la Iglesia G, Castro Solano A, Keyes CL. The Mental Health Continuum–Short Form (MHC–SF) in the Argentinean Context: Confirmatory Factor Analysis and Measurement Invariance. Europe’s Journal of Psychology. 2017 Mar 3; 13(1): 93–108.

20. The Copenhagen Burnout Inventory: A new tool for the assessment of burnout, Work and Stress 19(3): 192–207, July-September 2005

21. Wohl, MMJA et al., Looking Within: Measuring State Self-Forgiveness and Its relationship to Psychological Well-Being. Canadian Journal of Behavioral Science 2008, Vol 40(1): 1–10.

22. Bakker, A B et al., Using the jobs demands-resources model to predict burnout and performance. 2004 Human Resource Management, 43, 83–104.

23. Schaufeli WB, Toon TW. A Critical Review of the Job Demands-Resources Model: Implications for Improving Work and Health. From G.F. Bauer and O. Hämmig, Bridging Occupational, Organizational and Public Health: 43 A Transdisciplinary Approach, DOI 10.1007/978-94-007-5640-3_4,

24. Sestili C., Scalingi S., Cianfenilli S., et al., Reliability and Use of Copenhagen Burnout Inventory in Italian Sample of University Professors, Int J Environ Res Public Health. 2018 Aug; 15(8): 1708.

